# Prevalence and risk factors of neurodevelopmental disorders among migrant and refugee preschool children in high-income countries: A systematic review and meta-analysis protocol

**DOI:** 10.1101/2024.11.12.24317152

**Authors:** Kh Shafiur Rahaman, Valsamma Eapen, Mythily Subramanium, James Rufus John, Kanchana Ekanayake, Amit Arora

## Abstract

Neurodevelopment is a complicated mechanism involving genetic, cognitive, emotional, and behavioural processes. Factors related to parental migration directly or indirectly affect their children’s neurodevelopmental process and may lead to Neurodevelopmental Disorders (NDDs). Other factors such as barriers to accessing health services, social discrimination, mother’s psychosocial health during pregnancy may disrupt the neurodevelopmental process and lead to disorders and disabilities among children of migrants. However, there is a gap in data on the prevalence and the risk factors of neurodevelopmental disorders among migrant children have been inadequately listed. This paper presents a protocol for a systematic review to study and synthesise published evidence to ascertain the global prevalence of Neurodevelopment disorders and risk factors leading to those groups of neurodevelopment disorders among children of migrants in high-income countries. The protocol of this systematic review was developed with guidance from the Joanna Briggs Institute (JBI) methodology and reported as per the Preferred Reporting Items for Systematic Review and Meta-Analysis Protocols (PRISMA-P) statement. Observational studies that report on the prevalence and risk factors of neurodevelopment disorders among migrant young children under 5 years of age in high-income countries will be included in this study. Four electronic databases will be searched comprehensively (MEDLINE (Ovid), EMBASE (Ovid), CINAHL, PsycINFO, and Scopus). Two reviewers will independently screen, select studies, assess the methodological quality, and extract all relevant data subsequently. The systematic review and meta-analysis will help design tailored interventions for migrant children with and identify gaps from previous research to guide future research. This review is registered with PROSPERO (CRD42024589357).

## 1. Introduction

Neurodevelopmental disorders involve a group of conditions that start in the development phase of a child and disrupt the individual, socioeconomic, academic, and occupational functioning [1]. Neurodevelopmental Disorders broadly include intellectual disorders, autistic spectrum disorders (ASD), attention deficit hyperactivity disorders (ADHD), language and learning disorders, motor disorders, etc. [1]. Both genetic and environmental factors contribute to the development of these disorders [2], leading to impairments in motor, cognitive, language, and behavioural [3]. Globally, 316.8 million cases of developmental conditions have been reported, with males being affected more than females [3]. The prevalence of neurodevelopmental disorders among children aged between 0 to 18 years differs across different population groups. A recent systematic review reported a significant variation in the prevalence of neurodevelopmental disorders (4.7% to 88.5%) [4]. The prevalence of Neurodevelopment disorders in high-income countries was estimated between 7.0% to 15.0% of children in the general population [5, 6]. In contrast, the prevalence in low-middle countries has been reported to be 7.6% [7], and children from low-economic settings are asymmetrically affected by neurodevelopmental disorders [8].

Various risk factors of neurodevelopmental disorders have been reported in the literature, which include genetic, environmental, prenatal and postnatal issues, nutritional disorders, exposure to crisis, trauma, and poverty [9]. Furthermore, some children are more likely than others to develop developmental disabilities because of multiple unfavourable circumstances at different levels which include but are not limited to the family’s socioeconomic standing, their nationality, and their racial and ethnic heritage [3]. A recent review reported that children from migrant and refugee families coming from low and middle-income countries have a higher risk of neurodevelopmental disorders [10]. Children with one or both migrant parent(s) were found to have an increased risk of neurodevelopmental disorders related to language, academic skills, or coordination [11, 12].

The risk factors related to neurodevelopmental delays in children may vary based on parents’ ethnic background and circumstances during the migration [13, 14]. Other difficulties related to stigma [15], inadequate knowledge, differences in perception [16], referral patterns [17], and delayed diagnosis [18] also contribute to the risk of neurodevelopmental disorders among children with parental history of migration and ethnic minority. Migrants often exhibit risk factors such as inappropriate residence, inadequate nutrition (e.g., obesity) [19, 20], and low socioeconomic conditions [21], which are different from the host population. Maternal mental health is another risk factor for neurodevelopmental disorders. Some migrant mothers also would have experienced war and political conflict or extreme economic crises in their home country. Some mothers may have been exposed to poverty and sexual violence, which caused high mental stress during their pregnancy [22]. Health literacy among migrant families varies, as do the developmental outcomes of children. Additionally, cultural sensitivity and the capacity of health services in the host country to identify and treat developmental disabilities and impairments can differ significantly.

Available data on the prevalence and incidence rates of neurodevelopmental disorders are necessary to plan for the sustainable delivery of health, social and education services. However, data is often available only for the mainstream population, with only a few studies reporting on children with migrant and refugee backgrounds [25-27]. The main objective of this study is to conduct a systematic review and/or meta-analysis of the literature to explore the prevalence of neurodevelopmental disorders and their risk factors in young children (<6 years) with a parental history of migration in high-income countries. Specific review questions are the following:

i. What is the prevalence of neurodevelopmental disorders among preschool (<6 years) children of migrants, according to internationally published literature?
ii. What are the risk factors or associated factors of neurodevelopmental disorders among preschool (<6 years) children of migrants, according to internationally published literature?

## 2. Methods

### 2.1. Protocol and Registration

The systematic review will be conducted using the Joanna Briggs Institute (JBI) methodology for systematic reviews of effectiveness [28] and reported using the PRISMA (Preferred Reporting Items for Systematic Reviews and Meta-Analyses) guidelines [29]. The protocol for this review was reported as per the PRISMA for Systematic Review Protocols (PRISMA-P) framework. Additionally, the review has been registered with PROSPERO (CRD42024589357).

### 2.2 Review Question

The primary review questions were designed based on the population/participants, concept, and context (PCC) as recommended by the Joanna Briggs Institute JBI. The framework will suggest the search strategy and allow us to set the inclusion and exclusion criteria [30].

Among preschool children of migrant and refugee populations in high-income countries, what is the prevalence of neurodevelopmental disorders, and what are the risk factors affecting the disorders in question?

### 2.3 Inclusion Criteria

“Migrants and refugees” refer to groups of people moving for numerous reasons, often with overlapping motivations, as part of mixed movements – as defined by the United Nations High Commissioner for Refugees (UNHCR) [33]. According to the International Organization for Migration (IOM), a “migrant” is anyone who crosses an international border, leaving their birthplace or usual place of residence, for instance, unauthorised entrants, asylum seekers, and irregular or undocumented migrants. This applies regardless of their legal status and includes authorised migrants who left for work, family, and study [31]. Additionally, “refugee” encompasses individuals who have been granted refugee status of humanitarian protection and those fleeing persecution or organised violence – as defined by IOM [31]. In this study, children of migrants and refugee populations residing in high-income countries will be included, as classified by the World Bank (countries with a Gross National Income per capita of USD 13,205 or more) [32].

For our review, studies that included children aged between 0 and 5 years will be considered. Observational studies, such as cross-sectional and cohort studies (prospective and retrospective) that reported the prevalence of neurodevelopmental disorders and the risk factors among children of migrants, will also be included. Other studies, such as randomised controlled trials, case reports, opinions, letters to editors, and unpublished materials, will be excluded. Studies published in English in peer-reviewed journals will be included, with no restrictions on publication date.

### 2.4 Search Strategy

The search strategy will be developed with opinions from two expert health-sciences librarians. A mix of specific medical subject headings (MeSH), free-text terms, and Boolean operators will be included. The search terms will focus on prevalence, risk factors, migrants, refugees, preschool children, and relevant high-income countries. First, these terms will be tested in the Medline (OVID) database. The reviewers (K.S.R and A.A.) with experience in database searches will conduct an initial pilot search on two databases. The reviewers will then carry out all remaining literature searches independently. The preliminary Medline (OVID) search strategy formulated with guidance from expert librarians in the field is outlined in Table 2. After completing the Medline (OVID) database search, we will use the same syntax and subject headings for the remaining databases and adapt where necessary. We will manually review reference lists of the qualifying studies and existing literature reviews, including background and forward citation tracking of the studies included.

### 2.5 Information Sources

The following electronic databases will be searched without any restrictions on publication date (i.e., from the time of database inception to the present): MEDLINE (Ovid), Embase (Ovid), PsycINFO, Scopus, and Cumulative Index to Nursing and Allied Health Literature (CINAHL) (EBSCO). Search will be limited to English language only.

### 2.6 Study Selection

The studies identified through electronic databases and citations will be imported into EndNote 21 (Clarivate Analytics, Philadelphia, PA, USA). All the duplicates will be removed. After a pilot test, two independent reviewers (K.S.R. and A.A.) will review the titles and abstracts, adhering strictly to the inclusion and exclusion criteria. Total text will be retrieved for clarification if necessary. Then, the articles that meet the inclusion criteria will be retrieved in full, and their details will be imported into the JBI System for Unified Management, Assessment and Review of Information (JBI SUMARI) [34]. The same two reviewers will then independently evaluate the full texts to confirm eligibility. When we need additional information, the study authors will be contacted. Any disagreements with the reviewer will be addressed through discussion involving a third reviewer (J.J.) if necessary. The reasons for excluding full-text studies will be recorded and reported in the systematic review. If we find multiple articles published from the same study, they will be linked. The study selection process will be presented as a Preferred Reporting Items for Systematic Reviews and Meta-Analysis (PRISMA) flow diagrams [35].

### 2.7 Assessment of Methodological Quality

Each study included in this review will be assessed distinctly by two reviewers (K.S.R and A.A.) for methodological quality. A standardised critical appraisal tool developed by the Joanna Briggs Institute (JBI) will be used to determine the calibre of the articles included in the study [36]. Relevant quantitative tools will be used from JBI. Any disagreements will be solved between the two reviewers through discussion, and a third reviewer (J.J.) will be included when required. The authors of the study will be contacted to request missing or additional information where necessary to ensure the methodological quality. The studies will be assessed based on the available information if there is no response from the authors following two attempts. The risk of bias will be presented for each study through tables and figures and described narratively in the final review publication. The methodological concerns and their impact on result interpretation will be addressed in this narrative. All articles considered in this review will be subject to data extraction and synthesis, regardless of their methodological quality outcomes. Where possible, each study will be included for data extraction and synthesis irrespective of their methodological quality.

### 2.8 Data Extraction

A standard data extraction tool (see Table 1 in the Appendix) has been developed, which will be piloted in one study. The tool will be refined to ensure that all relevant data is captured. A calibration exercise will be conducted to ensure consistency among the two reviewers. Two authors (K.S.R and A.A.) will independently extract the discrepancies between them, which will be resolved by involving a third author (K.E.) when necessary. The extracted data will be recorded in an Excel spreadsheet with article details, participant characteristics, prevalence and risk factors of neurodevelopment disorders, type of migrant or refugee, study context, etc. Any additional information relevant to our study will be recorded, and the data extraction form will be modified accordingly. The authors of the study will be contacted when we require any additional information. If there is no response from the authors following two attempts, we will continue with the available information. The extracted data will be presented in narrative form and tables.

**Table 1.** Framework for inclusion criteria.

| Parameter |  | Criteria |
| --- | --- | --- |
| **P** | Population | Preschool children of migrants and refugees (as defined by the IOM (31), aged 0-6 years. |
| **C** | Concept | a. Prevalence of Neurodevelopmental Disorders (NDDs) as per the DSM-5<br>b. Risk factors/ predictors of NDDs |
| **C** | Context/ setting | High-income countries, as defined by the World Bank [32] |

**Table 2.**
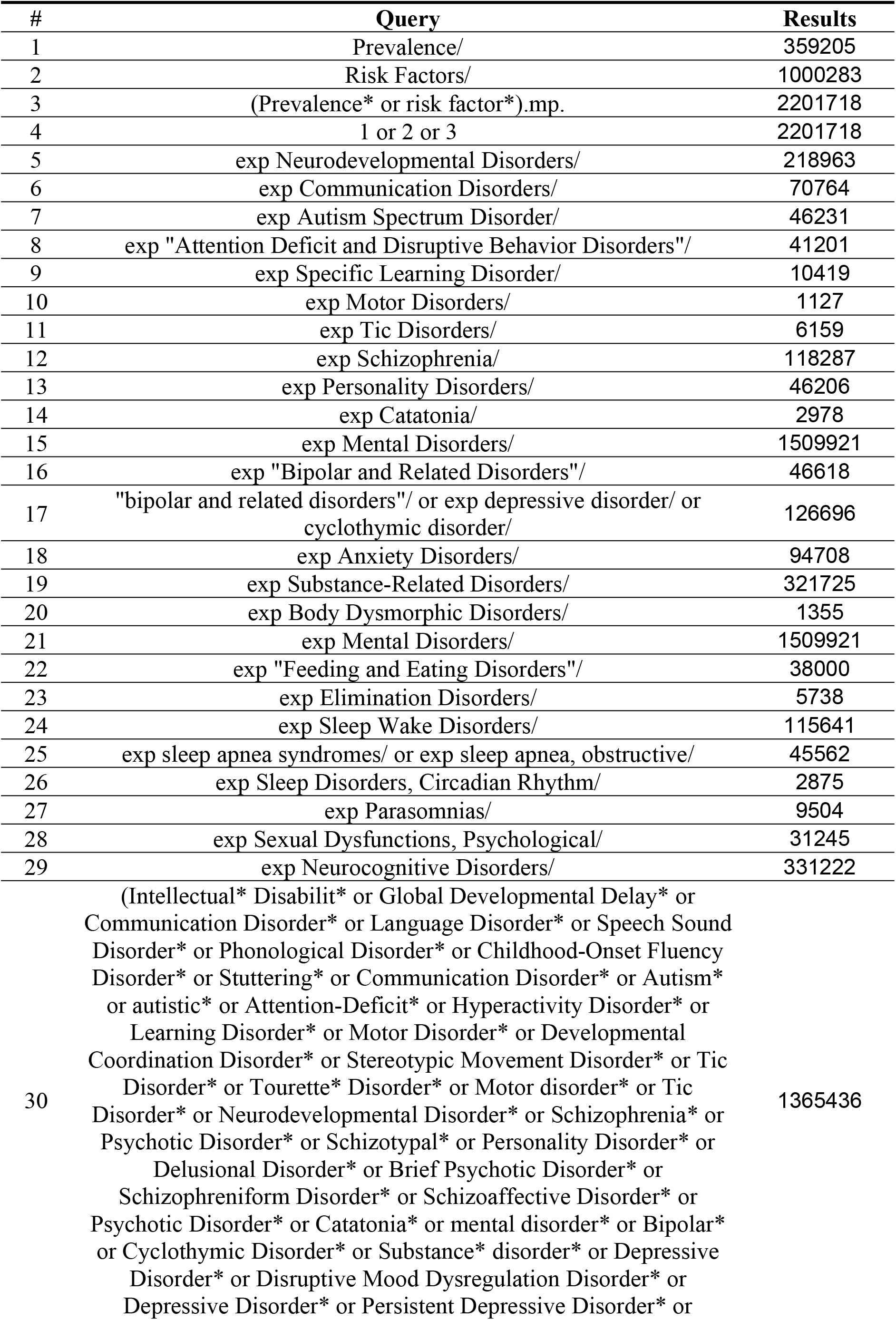
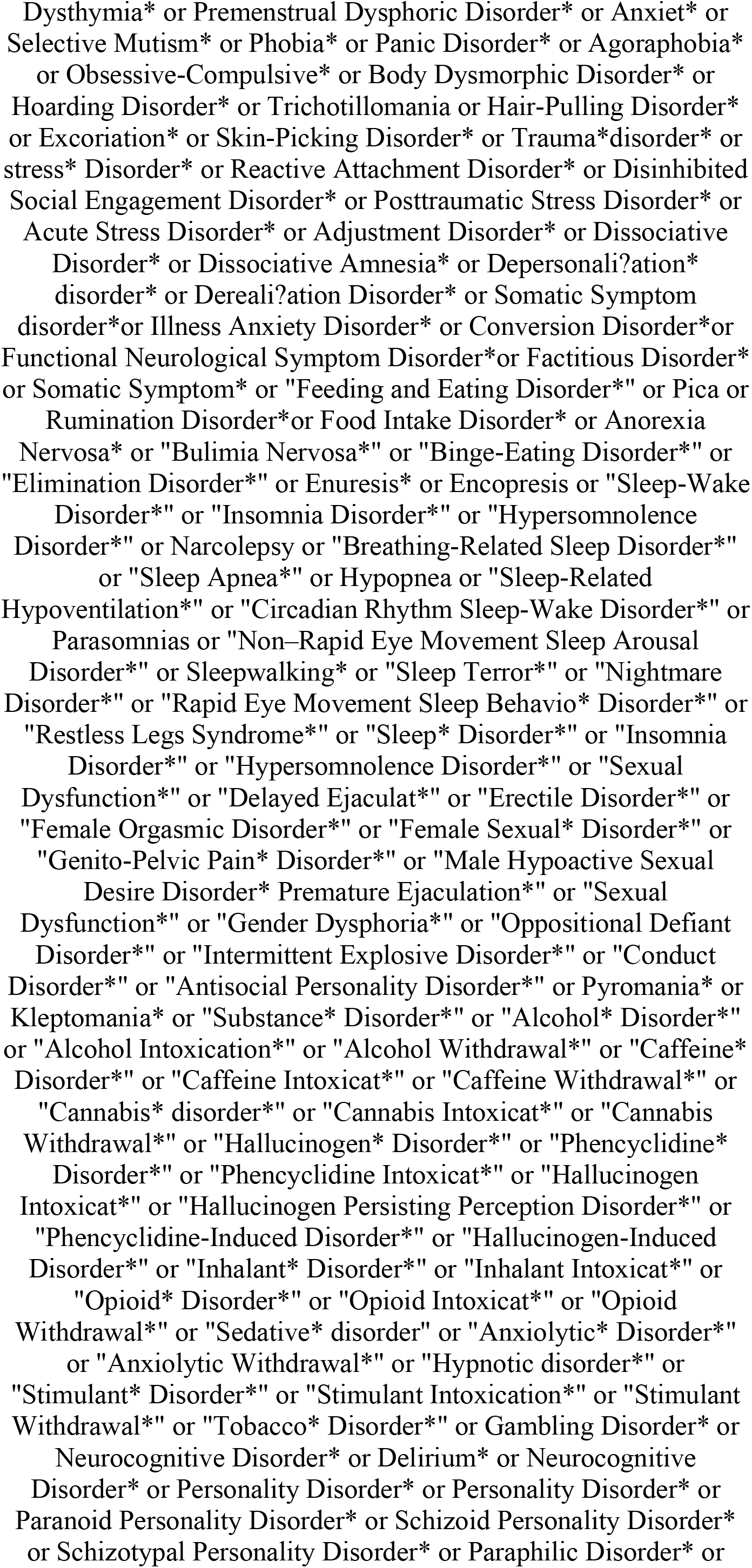
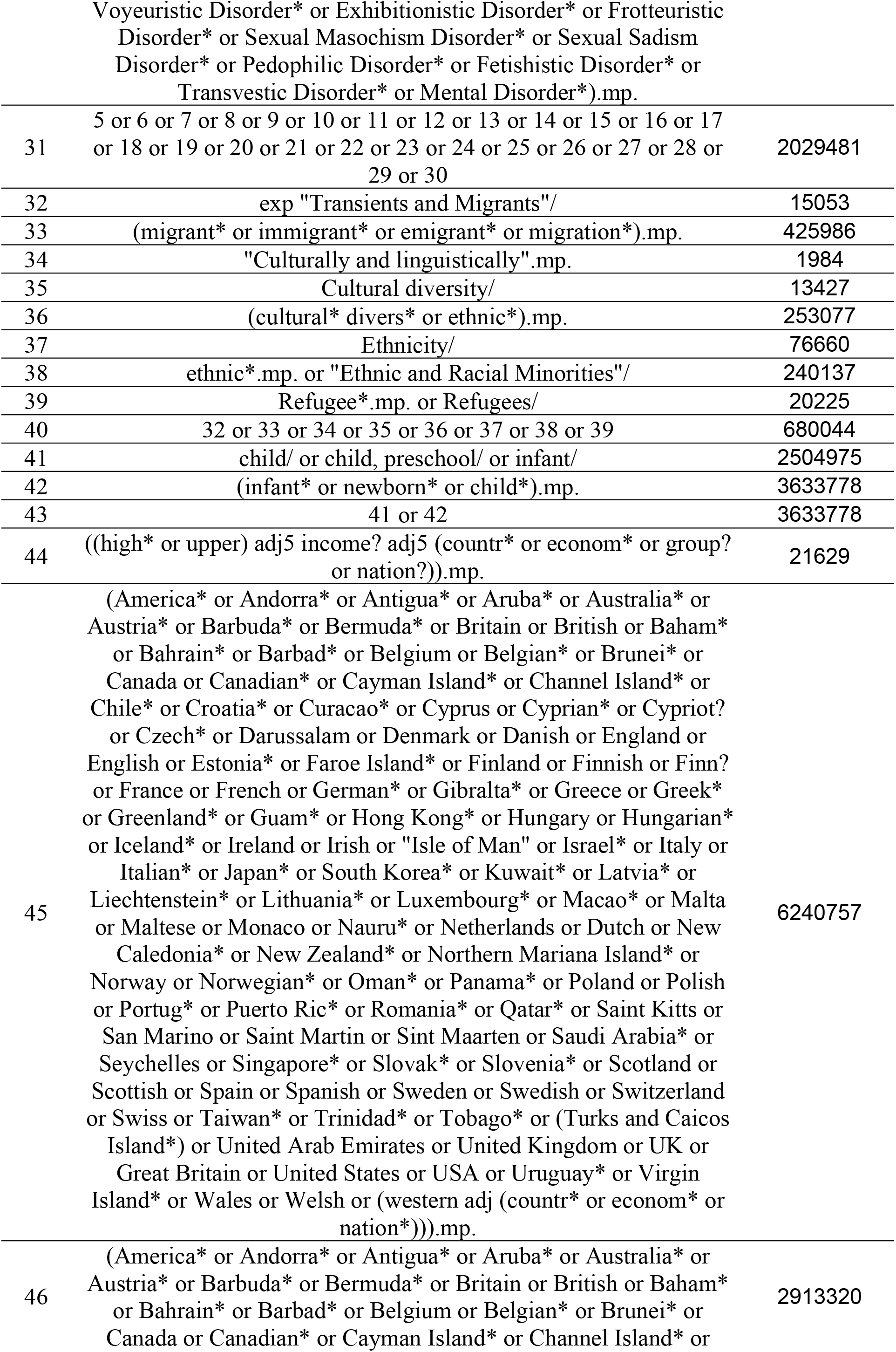
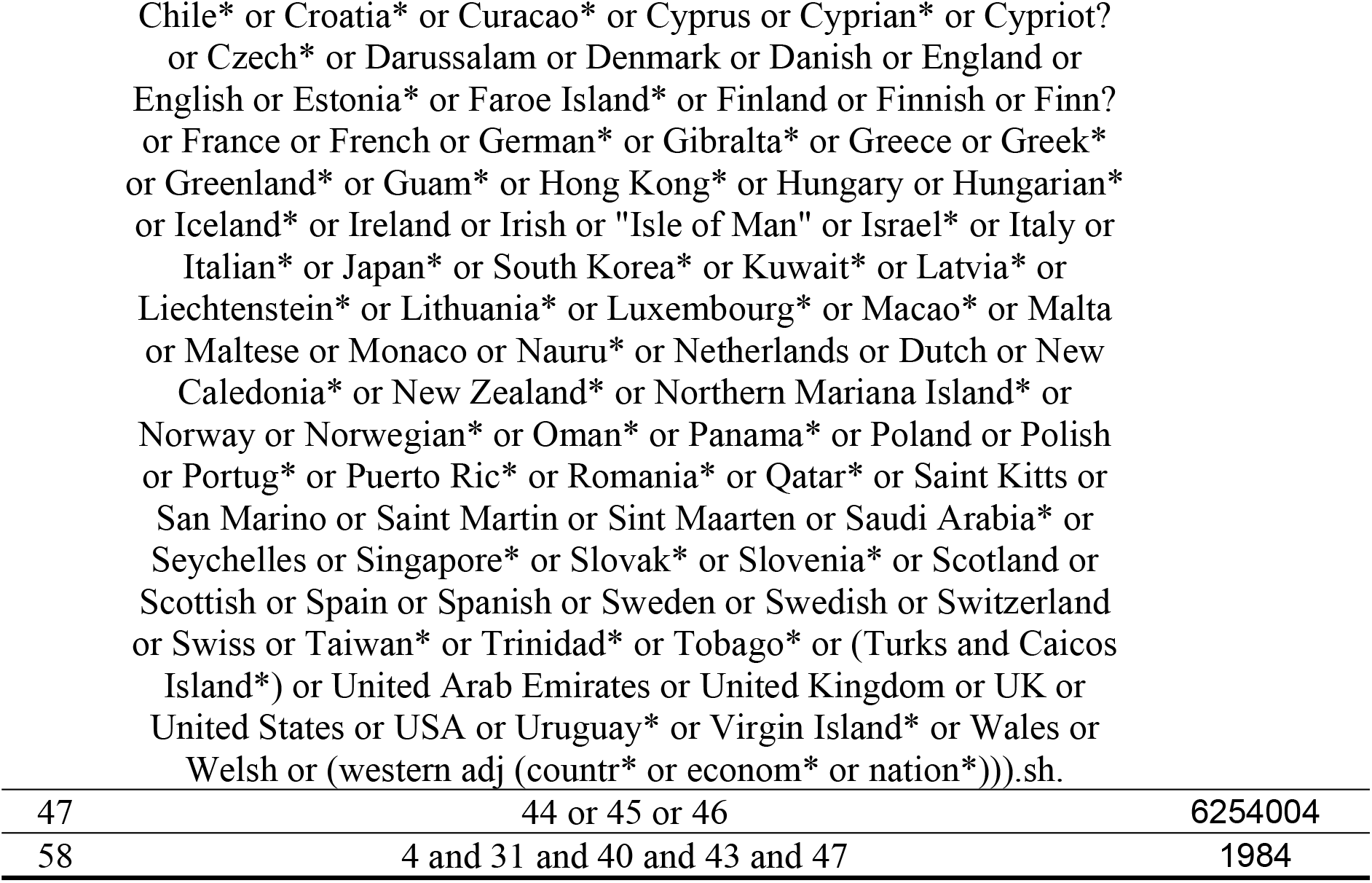
Primary search strategy for Medline (OVID) (2^nd^ of October 2024)

### 2.9 Data Synthesis

Quantitative data synthesis methods will be used to address the research findings comprehensively. The results will be categorised into settings, considering the origin of the participants and other environmental features. Depending on the number of eligible articles, a weighted inverse variance random-effects model will be used to estimate the overall pooled prevalence of neurodevelopmental disorders [37]. Otherwise, we will report the findings narratively. A subgroup analysis will be conducted based on the geographical area of the listed studies, study design, geographical origin of the study population, and type of neurodevelopment disorders to account for potential discrepancies in the overall prevalence estimates. Another subgroup analysis will be conducted in high-risk subgroups, such as recently resettled migrants/refugees (< 5 years versus long-term) and migrants/refugees from humanitarian crises. These subgroups could be more vulnerable to health conditions due to specific socioeconomic conditions, access to healthcare, and health inequalities. By identifying these sub-groups and analysing them separately, we aim to provide more unique insights into the comprehensive needs of these populations. Further sub-group analysis will be conducted based on the regional differences, for instance, population in North America, Europe, and Australia/New Zealand, to address contextual variability across high-income countries. Furthermore, a sensitivity analysis will be conducted to assess the robustness of the findings by excluding the studies with a high risk of bias or those that use different definitions or measurements of Neurodevelopment disorders. The publication bias among the studies will be assessed using a funnel plot and Egger’s regression test [38]. A narrative synthesis of the study findings will be used in tables and figures where statistical pooling is challenging due to heterogeneity among the studies. Stata 18 will be used for any statistical analysis to ensure rigorous data evaluation, leading to more valid and reliable conclusions.

## 3. Conclusions

The systematic review aims to provide overarching details on the prevalence of neurodevelopmental disorders and risk factors among preschool children of migrants (aged 0 to 6 years) in a global context. Understanding the risk factors is crucial to knowing what contributes to this population’s higher prevalence of neurodevelopmental disorders. The findings can help to design targeted interventions, develop culturally appropriate programs and attenuate these risks. Policymakers and health officials can use the review findings to address the unique challenges faced by children and their migrant parents within the health system. Our research also prioritises the necessity of including a diverse group in health research. Migrant population has their unique cultural practices, norms, and traditions. In this review, we will try to address this insufficiently represented group and contribute to equitable health research practices.

## Data Availability

No datasets were generated or analysed during the current study. All relevant data from this study will be made available upon study completion.

## Institutional Review Board Statement

Not applicable.

## Informed Consent Statement

Not applicable.

## Data Availability Statement

Not applicable.

## Author Contributions

K.S.R., K.E., and A.A. led the protocol design, search strategy, drafting, and revision of the manuscript. A.A. provided statistical advice and drafted and revised the manuscript. V.E., M.S., J.J., and A.A. were involved in the conception and design of the protocol, drafting, and revisions of the manuscript. All authors have read and agreed to the published version of the manuscript.

## Acknowledgements

This study is being completed as part of the Master of Research (MRes) degree for K.S.R. and acknowledges the Western Sydney University International Master of Research Scholarship.

## Appendix

**Table 1.** Data extraction template

| Study Details | Population | Setting | Country of Origin | Study Aims | Type of NDDs | Reported Prevalence | Statistical Methods and Risk factors | Article's conclusion/ Recommendations | Author's conclusion and Reviewer's comments |
| --- | --- | --- | --- | --- | --- | --- | --- | --- | --- |
| **Authors:**<br>*Authors et al. [ref]*<br>**Year of Publication:**<br>*Example: 2005*<br>**Study design:**<br>*Example: Cross-sectional study*<br>**Country:**<br>*Example: Australia* | **Sampling:**<br>Details on the number of participants and sampling technique.<br>**Participants details:**<br>Details on the characteristics of participants (age, sex, sociodemographic characteristics, etc.).<br>*Example:*<br>Participants included children of migrants | Details on the setting of the study.<br>*Example:*<br>The study was conducted in the City A in Country B, and among participants residing in middle income society. | Details on the country of origin of the participants family.<br>*Example:*<br>Participants parent(s)'s country of origin is Country X or Mother is from Country Y. | Details on the aims of the study. | Details on the type of NDDs<br>*Example:*<br>Autistic Spectrum Disorder | Details on the reported prevalence.<br>*Example:*<br>Prevalence of autism in this population was 8.5%. | Details on the statistical approach used in the study.<br>Example:<br>Regression analysis was used to predict factor Z.<br>**Risk factors:**<br>Details on the potential risk factor of disease A.<br>*Example:*<br>Variable X was a predictor or risk factor of disease X | Details on the Article's conclusion or Recommendations | Details on the **Author's conclusions.**<br>**Reviewer's comments:**<br>Review comments after evaluation of a publication, and brief explanation on its relation to the key research question of the systematic review.<br>**Assessment of methodological quality:**<br>This study has clearly met X/Y (x.x%) criteria in the critical appraisal tool. |

**Notes:** Explanation of certain concepts to provide clarity on the interpretation. *Example:* A, B, X and Y are the exemplar used for clarity.

## References

1. Psychiatric Association American. Neurodevelopmental disorders: DSM-5 selections. American Psychiatric Publishing; 2015.

2. Boivin MJ, Kakooza AM, Warf BC, Davidson LL, Grigorenko EL. Reducing neurodevelopmental disorders and disability through research and interventions. Nature. 2015 Nov 19;527(7578):S155–60. doi: 10.1038/nature16029. PMID: 26580321.

3. World Health Organization, United Nations Children’s Fund. Global report on children with developmental disabilities: from the margins to the mainstream. World Health Organization; 2023 Sep 15.

4. Frances L, Quintero J, Fernandez A, et al. Current state of knowledge on the prevalence of neurodevelopmental disorders in childhood according to the DSM-5: a systematic review in accordance with the PRISMA criteria. Child Adolesc Psychiatry Ment Health. 2022;16:27; 10.1186/s13034-022-00462-1

5. Halfon N, Houtrow A, Larson K, Newacheck PW. The changing landscape of disability in childhood. Future Child. 2012;22:13–42; 10.1353/foc.2012.0004

6. Boyle CA, Boulet S, Schieve LA, et al. Trends in the prevalence of developmental disabilities in US Children, 1997-2008. Pediatrics. 2011;127:1034–1042; 10.1542/peds.2010-2989.

7. Bitta M, Kariuki SM, Abubakar A, Newton CRJC. Burden of neurodevelopmental disorders in low and middle-income countries: a systematic review and meta-analysis. Wellcome Open Res. 2017;2:121–136; 10.12688/wellcomeopenres.13540.3

8. Durkin MS, Schneider H, Pathania VS, et al. Learning and Developmental Disabilities. In: Disease Control Priorities in Developing Countries. 2nd ed. The International Bank for Reconstruction and Development / The World Bank, Washington (DC); 2006. PMID: 21250359.

9. Durkin M. The epidemiology of developmental disabilities in low-income countries. Mental retardation and developmental disabilities research reviews. 2002;8(3):206–11.

10. Abdullahi I, Wong K, Mutch R, Glasson EJ, de Klerk N, Cherian S, Downs J, Leonard H. Risk of Developmental Disorders in Children of Immigrant Mothers: A Population-Based Data Linkage Evaluation. J Pediatr. 2019 Jan;204:275-284.e3. doi: 10.1016/j.jpeds.2018.08.047. Epub 2018 Oct 4. PMID: 30293641.

11. Loi S, Pitkänen J, Moustgaard H, Myrskylä M, Martikainen P. Health of immigrant children: the role of immigrant generation, exogamous family setting, and family material and social resources. Demography. 2021 Oct 1;58(5):1655–85.

12. Lehti V, Gyllenberg D, Suominen A, Sourander A. Finnish-born children of immigrants are more likely to be diagnosed with developmental disorders related to speech and language, academic skills and coordination. Acta paediatrica. 2018 Aug;107(8):1409–17.

13. Leonard, H., Glasson, E., Nassar, N., Whitehouse, A., Bebbington, A., Bourke, J., … & Stanley, F. (2011). Autism and intellectual disability are differentially related to sociodemographic background at birth. PloS one, 6(3), e17875.

14. Gerritsen AA, Bramsen I, Devillé W, van Willigen LH, Hovens JE, van der Ploeg HM. Physical and mental health of Afghan, Iranian and Somali asylum seekers and refugees living in the Netherlands. Soc Psychiatry Psychiatr Epidemiol. 2006 Jan;41(1):18–26. doi: 10.1007/s00127-005-0003-5. Epub 2006 Jan 1. PMID: 16341619.

15. Ikram UZ, Snijder MB, Fassaert TJ, Schene AH, Kunst AE, Stronks K. The contribution of perceived ethnic discrimination to the prevalence of depression. Eur J Public Health. 2015 Apr;25(2):243–8. doi: 10.1093/eurpub/cku180. Epub 2014 Nov 21. PMID: 25416918.

16. Ellen Selman L, Fox F, Aabe N, Turner K, Rai D, Redwood S. ‘You are labelled by your children’s disability’ - A community-based, participatory study of stigma among Somali parents of children with autism living in the United Kingdom. Ethn Health. 2018 Oct;23(7):781–796. doi: 10.1080/13557858.2017.1294663. Epub 2017 Mar 2. PMID: 28277014.

17. Fox F, Aabe N, Turner K, Redwood S, Rai D. “It was like walking without knowing where I was going”: A Qualitative Study of Autism in a UK Somali Migrant Community. J Autism Dev Disord. 2017 Feb;47(2):305–315. doi: 10.1007/s10803-016-2952-9. PMID: 27858263; PMCID: PMC5309314.

18. Begeer S, Bouk SE, Boussaid W, Terwogt MM, Koot HM. Underdiagnosis and referral bias of autism in ethnic minorities. J Autism Dev Disord. 2009 Jan;39(1):142–8. doi: 10.1007/s10803-008-0611-5. Epub 2008 Jul 4. PMID: 18600440.

19. Magaña S, Lopez K, Aguinaga A, Morton H. Access to diagnosis and treatment services among latino children with autism spectrum disorders. Intellect Dev Disabil. 2013 Jun;51(3):141–53. doi: 10.1352/1934-9556-51.3.141. PMID: 23834211.

20. Delobel-Ayoub M, Ehlinger V, Klapouszczak D, Maffre T, Raynaud JP, Delpierre C, Arnaud C. Socioeconomic Disparities and Prevalence of Autism Spectrum Disorders and Intellectual Disability. PLoS One. 2015 Nov 5;10(11):e0141964. doi: 10.1371/journal.pone.0141964. PMID: 26540408; PMCID: PMC4635003.

21. Miller LL, Gustafsson HC, Tipsord J, Song M, Nousen E, Dieckmann N, Nigg JT. Is the Association of ADHD with Socioeconomic Disadvantage Explained by Child Comorbid Externalizing Problems or Parent ADHD? J Abnorm Child Psychol. 2018 Jul;46(5):951–963. doi: 10.1007/s10802-017-0356-8. PMID: 29128953; PMCID: PMC5948120.

22. Ikram UZ, Snijder MB, Fassaert TJ, Schene AH, Kunst AE, Stronks K. The contribution of perceived ethnic discrimination to the prevalence of depression. Eur J Public Health. 2015 Apr;25(2):243–8. doi: 10.1093/eurpub/cku180. Epub 2014 Nov 21. PMID: 25416918.

23. Daseking M, Bauer A, Knievel J, Petermann F, Waldmann HC. Cognitive development and risk factors in bilingual pre-school children from an immigrant background. Praxis der kinderpsychologie und kinderpsychiatrie. 2011 Jan 1;60(5):351–68.

24. Graham HR, Minhas RS, Paxton G. Learning problems in children of refugee background: A systematic review. Pediatrics. 2016 Jun 1;137(6).

25. Crafa D, Warfa N. Maternal migration and autism risk: systematic analysis. International review of psychiatry. 2015 Jan 2;27(1):64–71.

26. Fairthorne J, de Klerk N, Leonard HM, Schieve LA, Yeargin-Allsopp M. Maternal race–ethnicity, immigrant status, country of birth, and the odds of a child with autism. Child neurology open. 2017 Jan 12;4:2329048X16688125.

27. Leonard H, Glasson E, Nassar N, Whitehouse A, Bebbington A, Bourke J, Jacoby P, Dixon G, Malacova E, Bower C, Stanley F. Autism and intellectual disability are differentially related to sociodemographic background at birth. PloS one. 2011 Mar 30;6(3):e17875.

28. Tufanaru C, Munn Z, Aromataris E, Campbell J, Hopp L. Systematic reviews of effectiveness. Joanna Briggs Institute reviewer’s manual. 2017 Apr 19;3.

29. Page MJ, McKenzie JE, Bossuyt PM, Boutron I, Hoffmann TC, Mulrow CD, Shamseer L, Tetzlaff JM, Akl EA, Brennan SE, Chou R. The PRISMA 2020 statement: an updated guideline for reporting systematic reviews. bmj. 2021 Mar 29;372.

30. Pollock D, Peters MD, Khalil H, McInerney P, Alexander L, Tricco AC, Evans C, de Moraes ÉB, Godfrey CM, Pieper D, Saran A. Recommendations for the extraction, analysis, and presentation of results in scoping reviews. JBI evidence synthesis. 2023 Mar 1;21(3):520–32.

31. International Organization for Migration. Glossary on Migration; International Organization for Migration (IOM): Geneva, Switzerland, 2019.

32. The World Bank. World Bank Country and Lending Groups; The World Bank: Washington, DC, USA, 2022.

33. Assembly UG. New York declaration for refugees and migrants. Resolution adopted by the General Assembly. 2016 Sep:19.

34. Munn Z, Aromataris E, Tufanaru C, Stern C, Porritt K, Farrow J, Lockwood C, Stephenson M, Moola S, Lizarondo L, McArthur A. The development of software to support multiple systematic review types: the Joanna Briggs Institute System for the Unified Management, Assessment and Review of Information (JBI SUMARI). JBI evidence implementation. 2019 Mar 1;17(1):36–43.

35. Page MJ, McKenzie JE, Bossuyt PM, Boutron I, Hoffmann TC, Mulrow CD, Shamseer L, Tetzlaff JM, Akl EA, Brennan SE, Chou R. The PRISMA 2020 statement: an updated guideline for reporting systematic reviews. bmj. 2021 Mar 29;372.

36. Munn Z, Moola S, Riitano D, Lisy K. The development of a critical appraisal tool for use in systematic reviews addressing questions of prevalence. International journal of health policy and management. 2014 Aug;3(3):123.

37. DerSimonian R, Kacker R. Random-effects model for meta-analysis of clinical trials: an update. Contemporary clinical trials. 2007 Feb 1;28(2):105–14.

38. Munn Z, Moola S, Riitano D, Lisy K. The development of a critical appraisal tool for use in systematic reviews addressing questions of prevalence. International journal of health policy and management. 2014 Aug;3(3):123.

